# A multidimensional cross-sectional analysis of COVID-19 seroprevalence among a police officer cohort: The PoliCOV-19 study

**DOI:** 10.1101/2021.08.09.21261789

**Authors:** Parham Sendi, Rossella Baldan, Marc Thierstein, Nadja Widmer, Peter Gowland, Brigitta Gahl, Annina Elisabeth Büchi, Dominik Güntensperger, Manon Wider, Manuel Raphael Blum, Caroline Tinguely, Cédric Maillat, Elitza S. Theel, Elie Berbari, Ronald Dijkman, Christoph Niederhauser

**Affiliations:** Institute for Infectious Diseases, University of Bern, Bern, Switzerland; Division Operations, Cantonal Police Bern, Bern, Switzerland; Interregional Blood Transfusion Swiss Red Cross, Bern, Switzerland; CTU Bern, University of Bern, Bern, Switzerland; Department of Emergency Medicine, Inselspital, Bern University Hospital, University of Bern, Bern, Switzerland; Department of General Internal Medicine, Inselspital, Bern University Hospital, University of Bern, Bern, Switzerland; Institute of Primary Health Care (BIHAM), University of Bern, Bern, Switzerland; Hôpital du Jura bernois SA, Saint-Imier, Switzerland; Division of Infectious Disease, Mayo Clinic, Rochester, MN, USA

## Abstract

**OBJECTIVE:** To determine the seroprevalence of SARS-CoV-2 antibodies in employees of the Cantonal Police Bern, Switzerland; to investigate individual and work-related factors associated with seropositivity; and to assess the neutralizing capacity of the antibodies of seropositive study participants.

**DESIGN:** Cross-sectional analysis of a cohort study.

**SETTING:** Wearing face masks was made mandatory for employees of the police during working hours at the rise of the second wave of the pandemic in mid-October 2020. Protests and police fieldwork provided a high exposure environment for SARS-CoV-2 infections. The investigation was performed prior to initiation of a vaccine programme. Study participants were invited for serological testing of SARS-CoV-2 and to complete questionnaires on sociodemographic, work and health-related questions.

**PARTICIPANTS:** 978 police personnel working in four different geographic districts, representing 35% of the entire staff, participated from February to March 2021.

**MAIN OUTCOME MEASURES:** Seroprevalence of anti-SARS-CoV-2 antibodies in February to March 2021, geographic and work-related risk factors for seropositivity, and serum neutralization titres towards the wild-type SARS-CoV-2 spike protein (expressing D614G) and the alpha and beta variants.

**RESULTS:** Seroprevalence was 12.9% (126 of 978 employees). It varied by geographic region within the canton; ranged from 9% to 13% in three regions, including the city; and was 22% in Bernese Seeland/Jura. Working in the latter region was associated with higher odds for seropositivity (odds ratio 2.38, 95% confidence interval 1.28 to 4.44, P=0.006). Job roles with mainly office activity were associated with a lower risk of seropositivity (0.33, 0.14 to 0.77, P=0.010). Most seropositive employees (67.5%) reported having had coronavirus disease 2019 (COVID-19) 3 months or longer prior to serological testing, and the proportion of agreement between positive nasopharyngeal test results and seroconversion was 95% to 97%. Among reported symptoms, new loss of smell or taste was the best discriminator for seropositivity (odds ratio 52.4, 30.9 to 89.0, P<0.001). Compliance with mask wearing during working hours was 100%, and 45% of all seropositive versus 5% of all seronegative participants (P<0.001) reported having had contact with a proven COVID-19 case living in the same household. The level of serum antibody titres correlated well with neutralization capacity. Antibodies derived from natural SARS-CoV-2 infection effectively neutralized the SARS-CoV-2 spike protein (expressing D614G), but were less effective against the alpha and beta variants. A regression model demonstrated that anti-spike antibodies had higher odds for neutralization than did anti-nucleocapsid protein antibodies.

**CONCLUSIONS:** Seroprevalence in the pre-vaccinated police cohort was similar to that reported in the general population living in the same region. The high compliance with mask wearing and the low proportion of seroconversion after contact with a presumed or proven COVID-19 case during working hours imply that personal protective equipment is effective and that household contacts are the leading transmission venues. The level of serum antibody titres, in particular that of anti-spike antibodies, correlated well with neutralization capacity. Low antibody titres were not effective against the alpha and beta variants.

**SUMMARY BOXES:** *WHAT IS ALREADY KNOWN ON THIS TOPIC:*

- The seroprevalence of anti-SARS-CoV-2 antibodies in the general population shows variations, depending on the geographic location of investigated study participants.
- Social distancing by avoiding crowds and maintaining a distance of 6 feet from others when in public are recommended. These recommendations are not realistic for security personnel and employees of a police department.
- Preventive strategies for individuals in a health care setting are warranted and effective to reduce potential exposures. The effect of preventive strategies on individuals working for the police force has not been investigated.

*WHAT THIS STUDY ADDS:*

- The study suggests that the overall seroprevalence of anti-SARS-CoV-2 antibodies among police officers is not higher than that in the general population, despite presumed higher exposure (e.g., public protests).
- Compliance with use of personal protective equipment among police officers was very high. The study results suggested that household contacts, rather than exposure during working hours, is the main source for viral transmission.
- Anti-SARS-CoV-2 antibodies derived from natural infection demonstrated good neutralization capacity towards strains that epidemiologically likely caused the infection, but moderate to poor neutralization capacity towards the alpha and beta variant.

## Introduction

Serological surveys that detect antibodies against SARS-CoV-2 antigens provide information on the prevalence in groups that might be more exposed to the virus or have had higher rates of infection.^1^ They help researchers to quantify the protective effect of mitigation efforts. Although the majority of workers exposed to proven SARS-CoV-2 are employed in healthcare sectors, other occupations have been associated with an increased risk for coronavirus disease 2019 (COVID-19). Police officers are one such commonly exposed group. In contrast to healthcare workers, this special population has contact with a frequently changing and unpredictable population.^2^ Physical distancing is often not possible. Unlike healthcare workers, police officers have no information pertaining to potential infectious diseases of the involved parties. Previous studies have shown that droplet and aerosol emission of contagious organisms occurs during speech^3-5^ and increases with voice loudness.^6^ These data underscore the risk of exposure to SARS-CoV-2 that may occur in fieldwork. This notion is even more important considering that the COVID-19 pandemic has ignited social unrest, including domestic violence and a surge in COVID-19 negations and anti-masking and anti-vaccine protests worldwide.^7-12^ It is reasonable to hypothesize that police officers, in particular those working in the field, are a high-exposure population.

To assess the risk for COVID-19 in this group, we are studying a cohort consisting of individuals employed by the Cantonal Police Bern in Switzerland.^13^ The aim of this cross-sectional analysis was to determine the seroprevalence of SARS-CoV-2 antibodies in employees of the cantonal police and to investigate individual and work-related factors associated with seropositivity. We also measured antibody titres of naturally acquired SARS-CoV-2 infection and correlated the results with the neutralizing capacity of the antibodies towards the “wild-type” SARS-CoV-2 spike (S) protein (expressing D614G) and the alpha and beta variants.

## Setting and Methods

### SARS-CoV-2 exposure

#### The first wave

The first case of COVID-19 was confirmed in Switzerland on February 25, 2020. On March 16, 2020, schools and most businesses were closed nationwide. On March 20, 2020, all gatherings of more than five people in public spaces were banned. The measures were gradually removed between late April and June 2020. On July 6, 2020, the Federal Council ordered mandatory face masks on public transport for all individuals 12 years of age or older.

#### The second wave

New measures were imposed in October 2020 as cases surged again.

#### Mask wearing

Wearing face masks for employees of the Cantonal Police Bern was recommended on August 28, 2020, and made mandatory during working hours on October 13, 2020. The types of mask provided by the police to their employees included surgical masks (type IIR) and police cloth masks certified by a material sciences and technology institute.^14^

#### Viral strains

The viral strain consisting of the mutation D614G in the S protein (Nextstrain clade 20A and its descendants) was the dominating circulating variant in 2020 in Switzerland.^15^ The alpha variant B.1.1.7 became the common variant in Switzerland starting mid-□February 2021.^16^

### Study population

The police force of the canton of Bern employs over 2,800 individuals, located across 58 police stations in cities and rural areas. The cantonal police have four regions of activity in bilingual (German and French) areas, including the capital of the country (Bern city). The departments are subcategorized as regional, criminal, prevention and environment, and others. Rules and regulations on hygiene precautions and mask wearing were identical for all districts.

### Recruitment period, questionnaires, and blood sampling

Study enrolment opened on December 21, 2020. Study participants who fulfilled the inclusion criteria (employees of the Cantonal Police Bern, aged 18-65 years) were asked to fill out two online questionnaires prior to being given appointments for blood sampling from February 9 to March 9, 2021. For every study participant, we obtained coded data on age and gender, education, job role within the police department, percentage employment, geographic region for work and living areas, and underlying health conditions. In addition, data on personal protective equipment use and hygiene precautions, symptoms consistent with COVID-19, contact with presumed or confirmed cases, quarantine and nasopharyngeal test results were obtained. The vast majority of nasopharyngeal swab tests included PCR technology, because rapid antigen tests became available through health professionals in Switzerland in November 2020 and for use by the general population in April 2021 (i.e., after recruiting and blood sampling). Answers to questions were provided prior to reporting the antibody test results.

After testing for the presence of antibodies, we contacted seropositive individuals again for an additional questionnaire on their subjective views about where the transmission possibly occurred.

### Antibody tests

SARS-CoV-2 antibodies to the nucleocapsid protein (NCP) and spike (S) protein were measured by using two commercially available immunoassays: anti-SARS-CoV-2 and anti-SARS-CoV-2 S (Roche Diagnostics, Rotkreuz, Switzerland). Both immunoassays detect antibodies independent of isotype, detecting predominantly IgG antibodies, as well as IgA and IgM antibodies. The anti⍰SARS⍰CoV⍰2 assay is based on antibody detection to a recombinant SARS-CoV-2 NCP, whereas the anti-SARS-CoV-2 S uses a recombinant receptor binding domain antigen, which is found on subunit 1 of the S protein. The NCP immunoassay reports a cut-off index (COI; signal of sample/cut-off) in which values ≥ 1.00 are considered positive, whereas the S immunoassay is a semi-quantitative method and reports results in absorbance units per millilitre (U/mL), for which values ≥ 0.8 AU/mL are considered positive. The tests were performed on the Roche cobas 8000 e801 analyser according to the manufacturer’s instructions (Roche Diagnostics, Rotkreuz, Switzerland).

### Test strategy for analysis

The cross-sectional baseline investigation was performed in a non-vaccinated population, because samples were obtained prior to initiating a vaccine programme for the employees of the police. First, all study participants were tested for the presence of anti-NCP antibodies.^17^ The rationale for this testing strategy included the high specificity of the anti-NCP electrochemiluminescence immunoassay (ECLIA) test evaluated in our laboratory.^18^ Second, all anti-NCP seropositive samples were then also tested for anti-S antibodies. The rationale for testing anti-S antibodies in NCP-seropositive individuals relied on the intended neutralization assays and correlation statistics between anti-NCP and anti-S antibody titres. Among seronegative samples, only those from individuals who reported having had a positive test result from a nasopharyngeal swab in the online questionnaire were tested for anti-S antibodies (**supplemental fig A**).

### Serum neutralization assays

The assays were performed as previously described.^19^ In brief, 20,000 Vero E6 cells were seeded in a 96-well plate format. The following day, heat-inactivated sera were two-fold serial diluted and mixed with 200 plaque forming units of the indicated isogenic SARS-CoV-2 virus, which were generated, rescued, and propagated.^19^ After 1 hour of pre-incubation at room temperature, the mixture was added to Vero E6 cells and incubated at 37°C. After 4 days, cells were fixed with 4% formalin and stained with crystal violet to analyse the reciprocal dilution at which SARS-CoV-2 was neutralized. We used isogenic SARS-CoV-2 viruses harbouring either the D614G spike, the full-length B.1.1.7 spike (alpha variant), or the full-length B.1.351 spike (beta variant).

### Statistical analysis

Seropositive individuals included those with a positive result and seronegative individuals those with a negative result in the anti-NCP antibody assay. The sample size calculation conducted prior to the study yielded a 95% confidence interval width of 4%, assuming a true seroprevalence of 12% to 13% and inclusion of 1,000 study participants.^13^ To describe characteristics of the study cohort, we used mean ± standard deviation (SD) or median with interquartile range for summarizing continuous variables, as appropriate. Comparisons were made by using the Student t test or Mann-Whitney test, respectively. Categorical data were shown as numbers with percentages and compared by using Fisher’s exact test for binary variables or the chi-squared test for more than two categories, unless indicated as having been tested for a non-parametric trend, following the approach of Cuzick.^20^ To investigate whether the outcome (seropositivity) was associated with a set of exposures, we used logistic regression on the entire cohort and on the sub-cohort of exposed subjects. We addressed the question of how well specific symptoms separate seropositive participants from those who tested negative by using random forests with 500 iterations. We calculated the odds ratio with the confidence interval, sensitivity, specificity, and C statistic (= area under the receiver operating characteristic curve) of the symptom with the highest importance. In seropositive patients, we visualized the correlation between antibody concentration (anti-NCP and anti-S) with maximal dilution of the neutralization tests and calculated Spearman correlation coefficients. We also investigated whether anti-NCP and anti-S antibody titres were equally associated with neutralization serum titres of the three different virus variants by using multilevel mixed effects ordered logistic regression (i.e., the level of serum neutralization titres as ordered categories). We calculated separate analyses, including both anti-NCP and anti-S antibodies as covariates into the model as continuous variables or dichotomized by the median, respectively. All analyses were performed with Stata 16 (Stata Corp., College Station, Texas).

## Results

A total of 989 individuals were enrolled in the study until March 9, 2021. Five individuals withdrew consent, and six missed the appointment for blood sampling. Hence, 978 employees of the Cantonal Police Bern were included in the final analysis, reflecting 35% of the entire staff. At the time of blood sampling, only two individuals were vaccinated with an mRNA vaccine (**supplemental fig A**). Both of them were anti-NCP seronegative.

### Seroprevalence

A total of 852 (87.1%) individuals were seronegative and 126 (12.9%) were seropositive at their anti-NCP antibody assay (**table 1**). Except for two individuals (1.6%), all anti-NCP seropositive samples were also positive for anti-S IgG (**supplemental fig A**).

**Table 1:** Demographic characteristics and anti-nucleocapsid protein IgG status for 978 employees of the Bern Cantonal Police.^1,2^

| Characteristics | N responses | Study participants (N=978) | Seropositive (N=126) | Seronegative (N=852) | P |
| --- | --- | --- | --- | --- | --- |
| **Age** – years; mean (SD) | 880 | 40 (8.9) | 39 (9.0) | 41 (8.8) | 0.12 |
| **Gender** – no. (%) | 973 |  |  |  | 0.67 |
| Female |  | 270 (28%) | 37 (29%) | 233 (27%) |  |
| Male |  | 703 (72%) | 89 (71%) | 614 (72%) |  |
| **Comorbidity** – no. (%) |  |  |  |  |  |
| Body mass index kg/m<sup>2</sup> | 975 | 26 (3.5) | 26 (3.3) | 26 (3.5) | 0.46 |
| Diabetes mellitus | 972 | 13 (1.3%) | 3 (2.4%) | 10 (1.2%) | 0.23 |
| Arterial hypertension | 973 | 78 (8.0%) | 10 (7.9%) | 68 (8.0%) | 1.00 |
| Cardiovascular disease | 971 | 17 (1.7%) | 3 (2.4%) | 14 (1.6%) | 0.47 |
| Lung disease | 973 | 27 (2.8%) | 2 (1.6%) | 25 (2.9%) | 0.56 |
| Immunosuppression | 971 | 12 (1.2%) | 2 (1.6%) | 10 (1.2%) | 0.66 |
| Other disease | 973 | 101 (10%) | 10 (7.9%) | 91 (11%) | 0.43 |
| No comorbidity | 978 | 761 (78%) | 100 (79%) | 661 (78%) | 0.73 |
| **Education** – no. (%) | 974 |  |  |  | 0.16 |
| Police academy |  | 795 (81%) | 114 (90%) | 681 (80%) |  |
| Security assistant school |  | 53 (5.4%) | 4 (3.2%) | 49 (5.8%) |  |
| University degree |  | 63 (6.4%) | 3 (2.4%) | 60 (7.0%) |  |
| Merchant |  | 32 (3.3%) | 2 (1.6%) | 30 (3.5%) |  |
| Craftsman |  | 18 (1.8%) | 2 (1.6%) | 16 (1.9%) |  |
| Other |  | 13 (1.3%) | 1 (0.79%) | 12 (1.4%) |  |
| **No. of years working for the police** – mean (SD) | 915 | 11 [7.0, 19] | 11 [7.0, 20] | 11 [6.5, 19] | 0.75 |
| **Working region within the canton** – no. (%) | 963 |  |  |  | **0.006** |
| Seeland, Bernese Jura |  | 170 (17%) | 37 (29%) | 133 (16%) |  |
| Mittelland, Emmental, Oberaargau |  | 193 (20%) | 22 (17%) | 171 (20%) |  |
| Bernese Oberland |  | 132 (13%) | 14 (11%) | 118 (14%) |  |
| Region Bern |  | 229 (23%) | 31 (25%) | 198 (23%) |  |
| Bern City |  | 239 (24%) | 22 (17%) | 217 (25%) |  |
| **Department** – no. (%) | 973 |  |  |  | **0.017** |
| Regional police |  | 649 (66%) | 90 (71%) | 559 (66%) |  |
| Criminal police |  | 118 (12%) | 20 (16%) | 98 (12%) |  |
| Traffic, environment, prevention |  | 72 (7.4%) | 10 (7.9%) | 62 (7.3%) |  |
| Interdepartmental |  | 120 (12%) | 6 (4.8%) | 114 (13%) |  |
| Other |  | 14 (1.4%) | 0 (0.00%) | 14 (1.6%) |  |
| **Main activity** – no. (%) | 939 |  |  |  | **0.038** |
| Fieldwork |  | 559 (57%) | 84 (67%) | 475 (56%) |  |
| Office work |  | 380 (39%) | 39 (31%) | 341 (40%) |  |
| % of working hours in the field, mean (SD) | 952 | 46 (27) | 51 (25) | 45 (27) | **0.024** |
| % of working hours in the office, mean (SD) | 976 | 50 [30, 80] | 40 [30, 70] | 50 [30, 80] | **0.012** |

<sup>1</sup>All except two individuals with anti-nucleocapsid protein (NCP) antibodies also displayed anti-spike antibodies (see **supplemental fig A**).
<sup>2</sup>Two vaccinated individuals (both anti-NCP antibody negative) were included in this analysis.

Seroprevalence varied by geographic region within the canton; it ranged from 9% to 13.5% in three regions, including the city, and was 22% in Bernese Seeland and Bernese Jura (**fig 1** and **table 1**).

**Figure 1:**
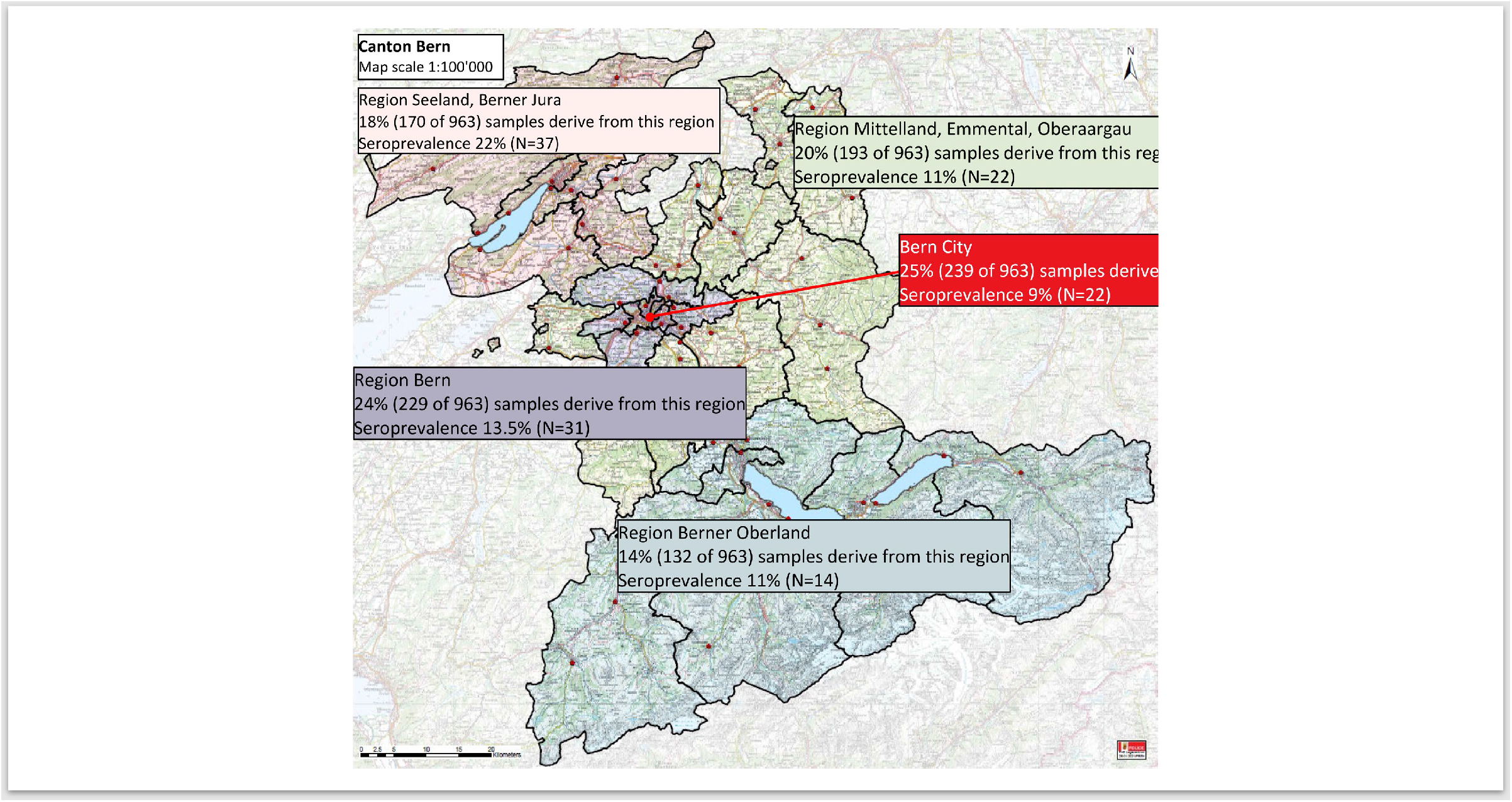
Map of the canton of Bern in Switzerland. The different colours indicate the corresponding geographic regions. The overall seroprevalence was 12.9% (i.e., 126 of 978 samples displayed anti-nucleocapsid protein antibodies). Responses of 963 study participants were available; 15 seronegative individuals did not provide their geographic working district.

Individuals who worked in the regions Bernese Seeland and Bernese Jura had significantly higher odds of having anti-SARS-CoV-2 antibodies than did those working in other regions (**supplemental table A**).

High public exposure (e.g., regional police with a high proportion of fieldwork activity) was associated with seropositivity (**table 1**). Conversely, roles with mainly office activity (i.e., interdepartmental) were associated with a lower risk of seropositivity (odds ratio 0.33, 95% confidence interval 0.14 to 0.77, P=0.010; **supplemental table B**).

In 85 (67.5%) of 126 seropositive individuals, the time interval between COVID-19 and blood sampling was 3 months or longer. There was no difference in anti-NCP and anti-S antibody titre results when we compared ECLIA values with the time intervals of 1 month, 2 months, 3 months, and longer (**supplemental fig B**).

Ninety-nine individuals reported having had a prior positive nasopharyngeal swab test result; 94 of them (95%) were seropositive for anti-NCP and 96 (97%) for anti-S antibodies. Only three (3%) of the positive tested individuals showed no seroconversion (**supplemental table C**). Three hundred thirty-six individuals reported never having had COVID-19 symptoms and that they were not tested; 329 (98%) of them were seronegative for anti-NCP antibodies.

### Symptoms

Symptoms consistent with COVID-19 were significantly more frequent in the seropositive than in the seronegative group (**table 2**). Among reported symptoms, “new loss of smell or taste” was the best discriminator (**supplemental fig C**). This symptom was associated with an odds ratio of 52.4 (95% confidence interval 30.9 to 89.0, P<0.001) for seropositivity, a sensitivity of 64% (55% to 72%), a specificity of 97% (95% to 98%), and an area under the receiver operating characteristic curve of 80% (76% to 85%).

**Table 2:**
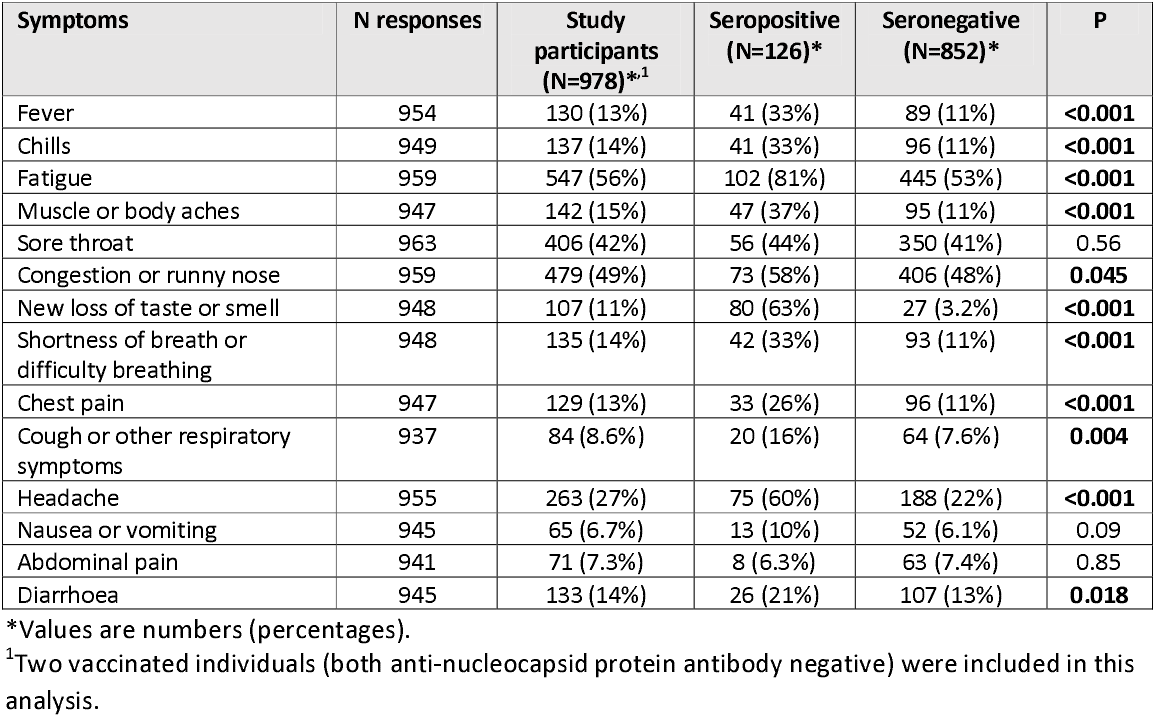
Clinical symptoms reported by study participants during the COVID-19 pandemic.

### Personal protective equipment use, absenteeism, quarantine, transmission venues

Reported compliance with wearing masks during working hours was 100%, irrespective of the presence of anti-SARS-CoV-2 antibodies (**supplemental table D**).

Two hundred forty-four individuals (25%) missed work (including home office) or were absent from the police academy because of suspected or confirmed COVID-19 or because of quarantine. Two hundred twenty-four (23%) individuals were placed in quarantine because of exposure to a person with proven or suspected COVID-19, and 65 (29%) were subsequently seropositive. These 65 employees represented 52% of all seropositive individuals in the study, while the other 159 employees who were placed in quarantine (and who remained seronegative) reflected 19% of all seronegative study participants (P<0.001). The investigation on possible transmission venues specifically differentiated between contacts during working hours and household, and between “presumed” and “proven” (i.e., confirmed with a nasopharyngeal swab test) COVID-19 contact. Fifty-seven (45%) of 126 seropositive individuals reported of having had contact with a proven COVID-19 case living in the same household. Conversely, 5% of all seronegative individuals reported the same type of exposure. This proportion difference was statistically significant (P<0.001). This was not the case when reported contacts during working hours or contacts with presumed (but not proven) cases were analysed (**table 3**).

**Table 3:**
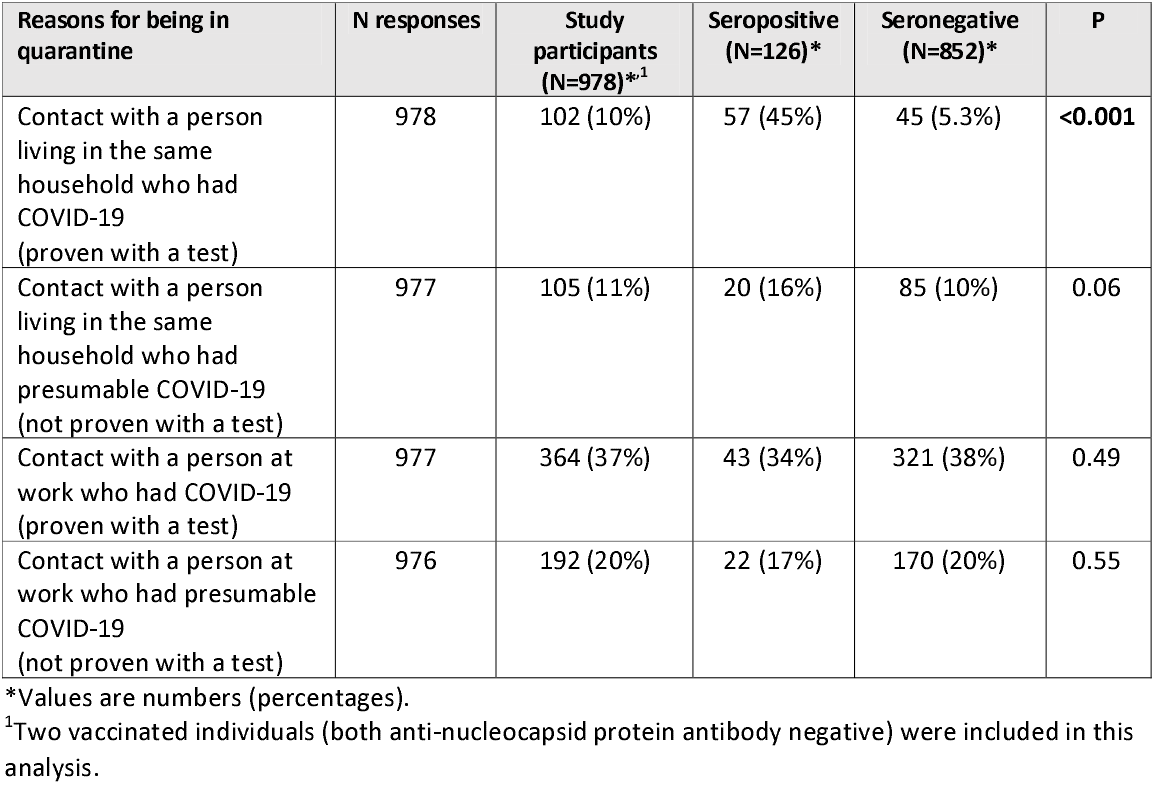
Comparison of seropositive (i.e., seroconversion) and seronegative (i.e., no seroconversion) individuals after having had contact with a proven or presumed COVID-19 case.

### Subjective assessment of study participants on the source of transmission

Among seropositive individuals, 75 (60%) felt “certain” or “likely certain” about the source of transmission (**supplemental table E**). Sixty-one (48%) reported that the contact had occurred during a private activity or within the same household.

### Neutralization capacity and correlation with ECLIA results

The required dilution titres for neutralization showed a considerable distribution among the study participants (**fig 2**). The median dilution titre was significantly higher in assays that used D614G than in those that used B.1.1.7 (P<0.001) and B.1.351 (P<0.001). Similarly, the median dilution titre was significantly higher in assays that used B.1.1.7 than in those that used B.1.351 (P<0.001). Antibody titres of anti-NCP- and anti-S antibodies correlated with the dilution titres showing the highest coefficient with dilution of D614G and the lowest coefficient with dilution of B.1.351 (beta), with P<0.001 in all pairs (**supplemental table F**). Antibodies from seropositive individuals demonstrated neutralization activity against D614G up to a dilution of 1:320 (**supplemental fig D**). The dilutions were lower for B.1.1.7 (1:40 for anti-S antibodies and 1:80 for anti-NCP antibodies) and for B.1.351 variants (1:20) (**supplemental figs E and F**), indicating a poorer neutralization capacity towards virus variants in comparison to the main circulating virus expressing D614G.

**Figure 2:**
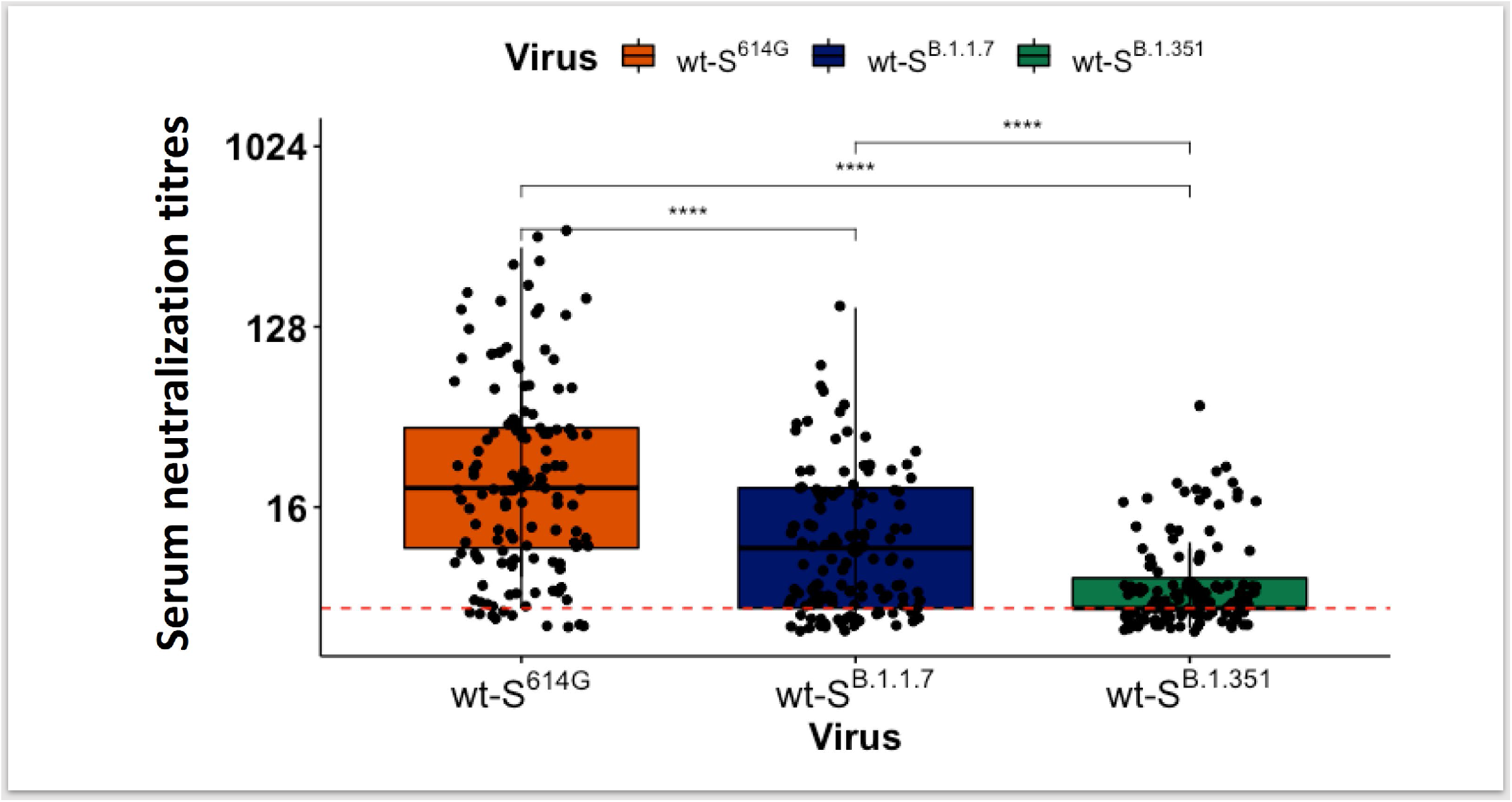
Results of neutralization assays performed with serum of study participants (n=126) and isogenic SARS-CoV-2 viruses harbouring either the D614G spike, the full-length B117 spike (alpha variant), or the full-length B.1.351 spike (beta variant). The red dashed line reflects the limit of detection. The numbers on the y-axis indicate the highest dilution of serum demonstrating neutralization activity. A Wilcoxon rank sum and signed rank test was performed to compare the groups. ^****^: P<0.0001. wt-S = wild-type spike.

We then searched for cut-off values of anti-NCP antibodies and anti-S antibodies in ECLIA results that demonstrated neutralization in the assay. We explored antibody titres as both continuous and binary variables in separate statistical models to mathematically predict the level of neutralization. For the latter, we used the median values of all results (i.e., >37.5 COI for anti-NCP antibodies and >65 U/mL for anti-S antibodies) and hence, assembled four combination categories (**fig 3**). Similar to the overall results of neutralization assays, the neutralization capacity of serum against alpha and beta variants was poorer than it was against D614G, even with serum demonstrating both >37.5 U/mL COI anti-NCP antibodies and >65 U/mL anti-S antibodies. In the model with both continuous and ordered categories, odds ratios of anti-S antibodies for the level of neutralization were higher than those of anti-NCP antibodies. While the antibody titres above the cut-off level of >37.5 U/mL were associated with about a three-fold increase in level of neutralization, anti-S1 antibody titres above >65 U/mL were associated with about a six-fold increase (**supplemental table G**).

**Figure 3:**
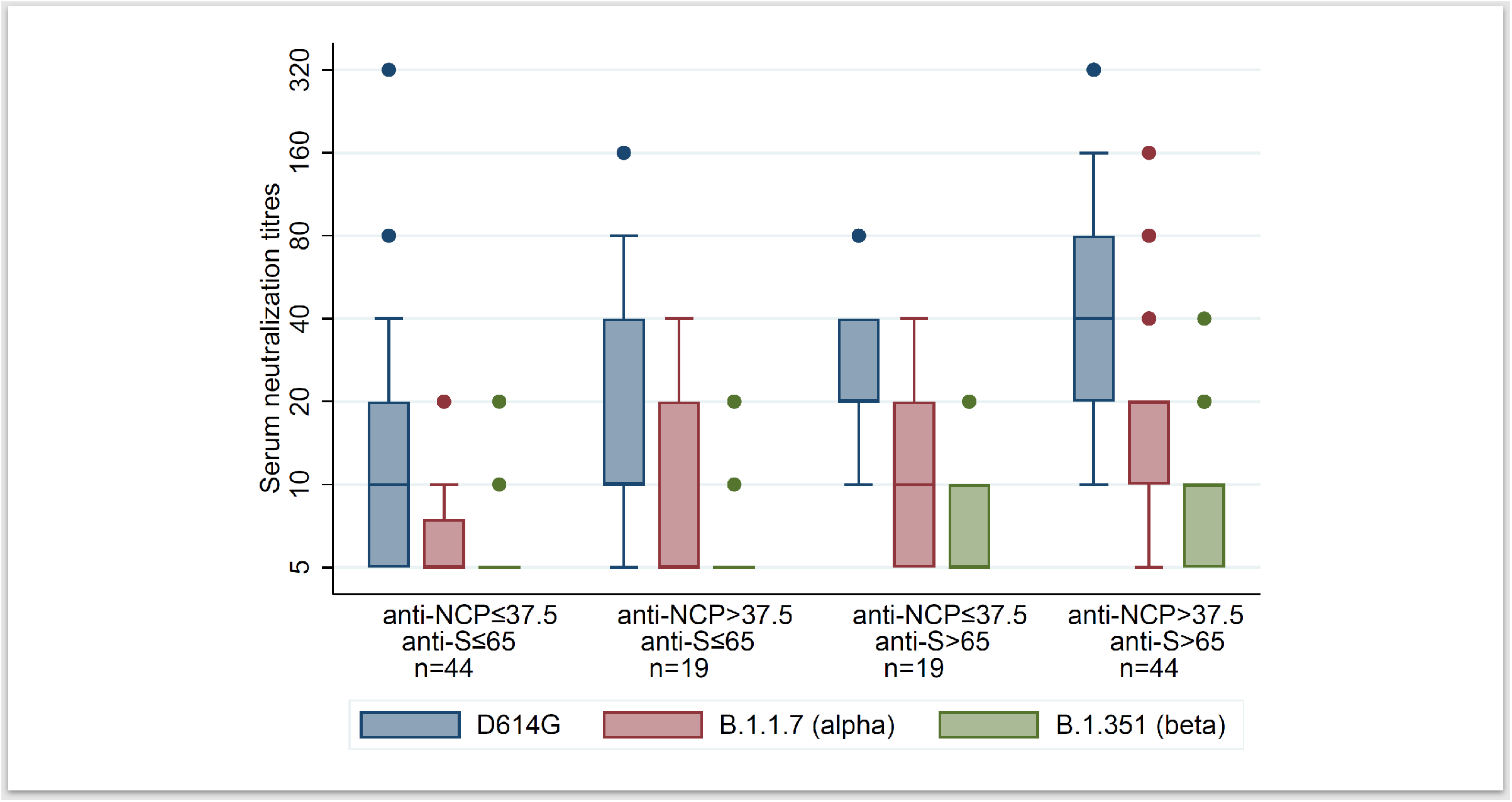
Results of neutralization assays categorized according to selected cut-off ELISA values of anti-nucleocapsid protein (NCP) antibodies (cut-off index ≤37.5 and >37.5) and anti-spike (S) antibodies (≤65 U/mL and >65 U/mL). The numbers on the y-axis indicate the highest serum dilution demonstrating neutralization activity.

## Discussion

This cross-sectional population serological survey among the police cohort demonstrated a pre-vaccinated anti-SARS-CoV-2 antibody seroprevalence of 13%. Few studies have investigated seroprevalence in police officers, which include those performed in New York, NY, USA, **^21^** and Mazowieckie Province, Poland, **^22^** as well as two further studies with very low sample sizes. **^23, 24^**However, our study did not demonstrate a higher seroprevalence than observed in the general population of the canton of Bern (i.e., 14%), which was investigated in another study using a different serological test.^25 26^ The results indicate that the use of personal protective equipment is effective in mitigating the risk of COVID-19. This is in line with the reported high compliance with mask wearing in our study population. However, within the police cohort, the odds for seropositivity were higher for fieldwork activity with high exposure to the general population than they were for office work activity with low exposure to the general population. The significant seroprevalence difference observed for contacts in the private environment and when comparing geographic districts is in line with the findings of others.^21 27 28^

During the first wave of the pandemic in Switzerland, testing individuals with few or no symptoms was not recommended, and symptoms consistent with COVID-19 were presumed to be COVID-19 related. In the police cohort, nearly 50% of individuals experienced sore throat, congestion, or a runny nose, although only 5% to 7% were seropositive. In our analysis, only new loss of taste or smell was associated with high specificity in predicting seropositivity, underscoring the importance of testing, considering that numerous other viruses can cause a variety of respiratory symptoms. Following proven infection with SARS-CoV-2, most individuals in our cohort developed detectable serum antibodies towards both the NCP and the S protein. To assess humoral immunity within our cohort, we investigated the magnitude of neutralizing antibodies towards SARS-CoV-2 strains that did and did not circulate within the population prior to blood sampling.

Considering that the sampling occurred from February 9 to March 9, the vast majority of our cohort was exposed to the virus strain harbouring the D614G S protein.^15^ Exposure to the alpha and beta variant was unlikely in our cohort,^16^ in particular when considering he reported time points of infection (**supplemental fig B**). In line with these observations, naturally acquired antibodies demonstrated good neutralization activity against D614G but performed suboptimally against the alpha variant and poorly against the beta variant. Understandably, we observed a correlation between the ECLIA titres and the highest dilutions still demonstrating neutralization. Our results imply that the interaction between the S protein and anti-S antibodies may play an important role in the neutralization tests. They point towards the importance of high anti-S antibody titres, and hence, the value of the vaccine achieving this goal. Although the variants show mutations mainly in the S protein, variations in the capsule have gained less attention. In line with our results, the contribution to virus neutralization of anti-NCP antibodies is less known.^29^

Our study has limitations. The investigation was performed in February 2021 and we used anti-NCP antibodies as the main marker of seropositivity. We cannot exclude that in certain individuals with COVID-19 in 2020 anti-NCP antibodies have waned below the detection level and that the true seroprevalence is underestimated. In our experience, the antibody titres remain at a detectable level for a prolonged period, and the proportion of agreement with reported nasopharyngeal swab test results was high. Therefore, and in consideration of a 95% confidence interval width of 4%, we are convinced that the seroprevalence proportion found in our analysis is a valid result. We categorized our analysis only in anti-NCP seropositive and seronegative individuals and did not correct for sensitivity and specificity. It is therefore possible that we included very few false positive or false negative serum samples in our analysis. Given the high sensitivity and specificity of the antibody tests, we do not believe that excluding these few samples would have changed the overall results. The results from nasopharyngeal swab tests were obtained via online questionnaire, and the questionnaires on the subjective view of transmission routes may consist of a recall bias. The choice of median cut-off values of antibody titres in association with serum neutralization assays is arbitrary and does not reflect clinical circumstances. More sophisticated methods to find an optimal cut-off such as receiver operating characteristic curves would have required a fixed and already established cut-off for serum neutralization titre. We chose a conservative cut-off, supported by the sensitivity analysis with antibody titres as a continuous independent variable.

In conclusion, our COVID-19 cross-sectional survey among police officers demonstrated a seroprevalence of 13% in a pre-vaccinated cohort. This proportion is similar to that reported in the general population. The high compliance with mask wearing and the low proportion of seroconversion after contact with a presumed or proven COVID-19 case during working hours imply that personal protective equipment is effective. The high proportion of seropositive individuals who have had contact with a proven COVID-19 case in the same household indicates that most transmissions within our police cohort did not occur within working hours. The level of serum antibody titres, in particular that of anti-S antibodies, correlated well with the neutralization capacity. Antibodies derived from natural SARS-CoV-2 antibodies effectively neutralized viral strains that – from an epidemiological point of view – most likely caused the infection. However, at low titres (i.e., below the median of the study population), antibodies were not effective against the alpha and beta variants. These findings may support decisions of policy makers and vaccine initiators to maintain public protective services on a stably staffed level.

## Data Availability

Requests for additional data and analytic scripts used in this study should be emailed to PS.

## Acknowledgment

We thank Dr. Volker Thiel (Institute of Virology and Immunology and Department of Infectious Diseases and Pathobiology, Vetsuisse Faculty, University of Bern, Bern, Switzerland) for providing the viral strains. We thank the numerous volunteers and the study team members from the affiliated and other institutions for their help in conducting the study, including Medbase Zweisimmen, Medicentre Moutier, and numerous others. We thank all the employees of the Cantonal Police Bern for participating. We are grateful to Annetta Redmann, CTU, University of Bern, for her databank and data management. BioMedical Editor, St Albert, Alberta, Canada, provided English-language editing.

## Contributors

PS, RB, and CN designed the study and contributed to the protocol. MT was responsible for conducting the study within the police departments. NW, PG, and CN were responsible for performing the ECLIA tests and the secure handling and archiving of serum samples. DG and BG managed the data. BG performed independent analyses in R and Stata. AEB was responsible for data monitoring. MB provided his expertise in epidemiology and participated in the field study. MW and RD were responsible for performing neutralization assays and interpreting the results. CM contributed to the coordination and conduction of the study in the Bernese Jura. PS, RB, CN, PG, and RD drafted the initial manuscript. EB and ET reviewed the initial draft and provided their scientific expertise in COVID-19. PS provided clinical interpretation and all authors contributed to the revision of the manuscript. PS is the guarantor. The corresponding author attests that all listed authors meet authorship criteria and that no others meeting the criteria have been omitted.

## Competing interest

All authors have completed the ICMJE uniform disclosure form at www.icmje.org/coi_disclosure.pdf and declare no conflict of interest.

Roche (manufacturer of the ECLIA tests) had no influence in any part of this study. The test was commercially purchased by the investigators. The lead author (PS) affirms that the manuscript is an honest, accurate, and transparent account of the study being reported and that no important aspects of the study have been omitted.

## Ethical approval

All participants gave written informed consent before participation in the PoliCOV-19 study.

The study was approved by the Cantonal Research Ethics Commission of Bern, Switzerland (ID-2020-02650).

## Data sharing

Requests for additional data and analytic scripts used in this study should be emailed to PS. This is an Open Access article distributed in accordance with the Creative Commons Attribution Non Commercial (CC BY-NC 4.0) license, which permits others to distribute, remix, adapt, and build upon this work non-commercially, and to license their derivative works on different terms, provided the original work is properly cited and the use is non-commercial. See: http://creativecommons.org/licenses/by-nc/4.0/.

## Dissemination to participants and related patient and public communities

This topic has garnered interest from academia, policy makers, social commentators, and many parts of the media. We will disseminate the results by using traditional academic publications such as this, traditional media through our media offices, and contemporary social media platforms.

