## Supplemental Material for "A multidimensional cross-sectional analysis of COVID-19 seroprevalence among a police officer cohort: The PoliCOV-19 study"

| Content | Page |
|---|---|
| **Supplemental figure A:** Antibody test results according to the test strategy. | 2 |
| **Supplemental table A**: Association of working region and the presence of anti-NCP antibodies. | 2 |
| **Supplemental table B**: Association of working department and the presence of anti-NCP antibodies. | 2 |
| **Supplemental table C:** Comparison between seropositive and seronegative individuals categorized according to the reported test results from nasopharyngeal swabs or COVID-19 symptoms prior to February 2021. | 3 |
| **Supplemental figure B:** Antibody titres of seropositive study participants in association with the time point of reported test results from nasopharyngeal swabs. | 3 |
| **Supplemental figure C:** Random forest algorithm to identify specific symptoms that separated seropositive and negative participants. | 4 |
| **Supplemental table D:** Comparison between seropositive and seronegative individuals categorized according to the use of personal protective equipment and hygiene precautions. | 4 |
| **Supplemental table E:** Subjective assessment of the source of transmission in seropositive study participants. | 5 |
| **Supplemental table F:** Spearman's rank correlation coefficients between serum antibody titres of anti-NCP and anti-S antibodies and serum neutralization titres towards SARS-CoV-2. | 6 |
| **Supplemental figure D:** Antibody titres of seropositive study participants in association with dilution titres in neutralization assays against isogenic SARS-CoV-2 viruses harbouring the D614G spike. | 6 |
| **Supplemental figure E:** Antibody titres of seropositive study participants in association with dilution titres in neutralization assays against isogenic SARS-CoV-2 viruses harbouring the full-length B.1.1.7 spike. | 7 |
| **Supplemental figure F:** Antibody titres of seropositive study participants in association with dilution titres in neutralization assays against isogenic SARS-CoV-2 viruses harbouring the full-length B.1.351 spike. | 7 |
| **Supplemental table G:** Association of antibody titres (≤37.5 and >37.5 COI for anti-NCP-antibodies and ≤65 and >65 for anti-S antibodies) and serum neutralization titres. | 8 |

**Supplemental figure A:** Antibody test results according to the test strategy.

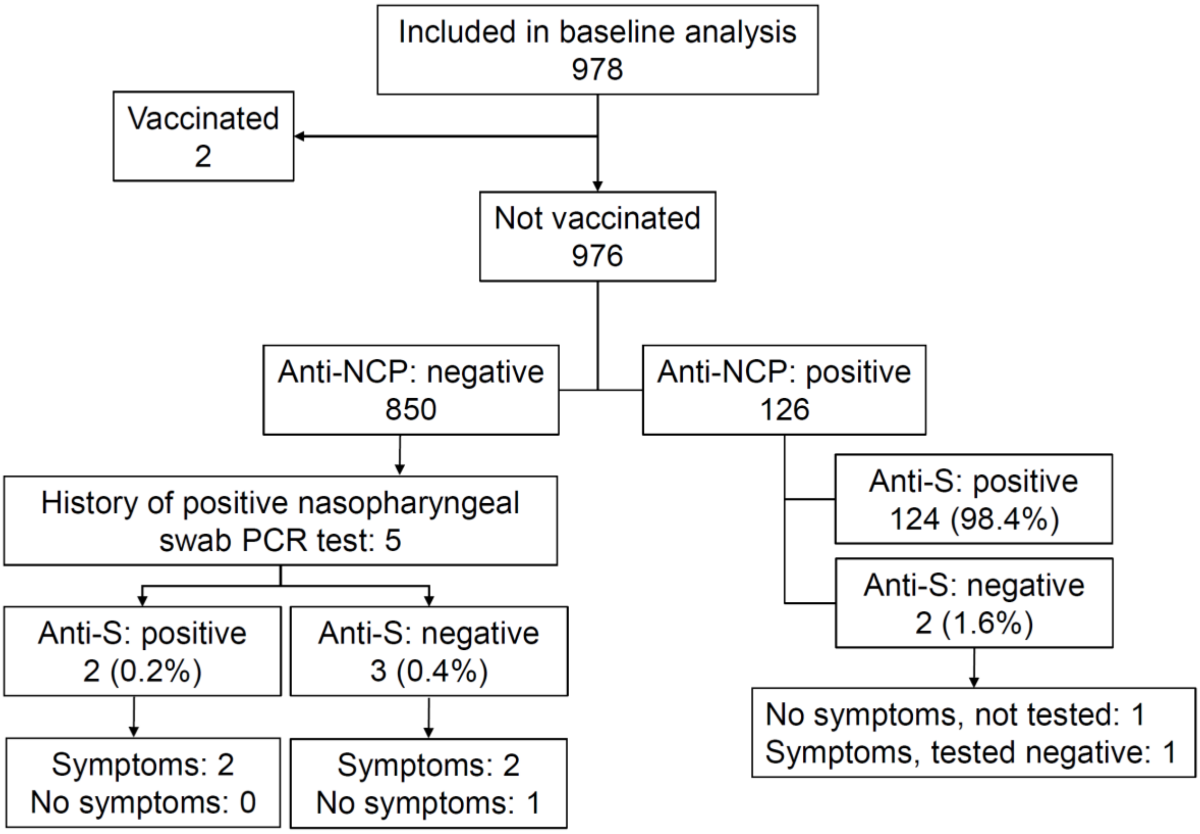


"Tested negative" (right bottom box) refers to reported nasopharyngeal swab test. The two vaccinated individuals were anti-NCP negative, one anti-S seropositive and one anti-S seronegative.

**Supplemental table A**: Association of working region and the presence of anti-NCP antibodies.

| **Geographic working region** | **OR (95% CI)** | **P value** |
|---|---|---|
| Bern City | Reference | |
| Region Bern | 1.57 (0.83 to 2.98) | 0.163 |
| Mittelland, Emmental, Oberaargau | 1.09 (0.56 to 2.14) | 0.799 |
| Bernese Oberland | 0.99 (0.47 to 2.10) | 0.987 |
| **Bernese Seeland, Bernese Jura** | **2.38 (1.28 to 4.44)** | **0.006** |

N=978

**Supplemental table B**: Association of working department and the presence of anti-NCP antibodies.

| **Department** | **OR (95% CI)** | **P value** |
|---|---|---|
| Regional police | Reference | |
| Criminal police | 1.27 (0.75 to 2.15) | 0.381 |
| Department of traffic, environment, prevention | 1.00 (0.50 to 2.03) | 0.996 |
| **Interdepartmental** | **0.33 (0.14 to 0.77)** | **0.010** |
| Others* | - | - |

N=978; *not estimable due to zero events.

**Supplemental table C:** Comparison between seropositive and seronegative individuals categorized according to the reported test results from nasopharyngeal swabs or COVID-19 symptoms prior to February 2021.

| **COVID-19 symptoms and test results from nasopharyngeal swabs** | **Study participants N=978** | **Seropositive** Anti-NCP | **Seronegative** |
|---|---|---|---|
| Never had symptoms, was not tested | 336 (34%) | 7 (5.6%)[1] | 329 (39%) |
| Had symptoms but was not tested | 139 (14%) | 9 (7.1%) | 130 (15%) |
| Had symptoms or contact but tested negative (nasopharyngeal swab) | 398 (41%) | 16 (13%)[2] | 382 (45%) |
| Had no symptoms but tested positive in nasopharyngeal swab test (e.g., contact tracing) | 6 (0.61%) | 5 (4.0%)[3] | 1 (0.12%) |
| Had symptoms and tested positive in nasopharyngeal swab test | 93 (10%) | 89 (71%)/ 91 (72%)[4] | 2 (0.2%)[4] |

N = 972 responses; the distribution of responses and results was significant (P<0.001).

[1] One of the seven anti-NCP seropositive individuals did not display anti-S antibodies in the ECLIA test.

[2] One of the 16 anti-NCP seropositive individuals did not display anti-S antibodies in the ECLIA test.

[3] All five anti-NCP seropositive individuals also displayed anti-S antibodies in the ECLIA test.

[4] Two anti-NCP seronegative individuals displayed positive anti-S antibodies in the ECLIA test.

**Supplemental figure B:** Antibody titres of seropositive study participants (blood sampling from February 9 to March 9, 2021) in association with the time point of reported test results from nasopharyngeal swabs (x-axis); (N=126).

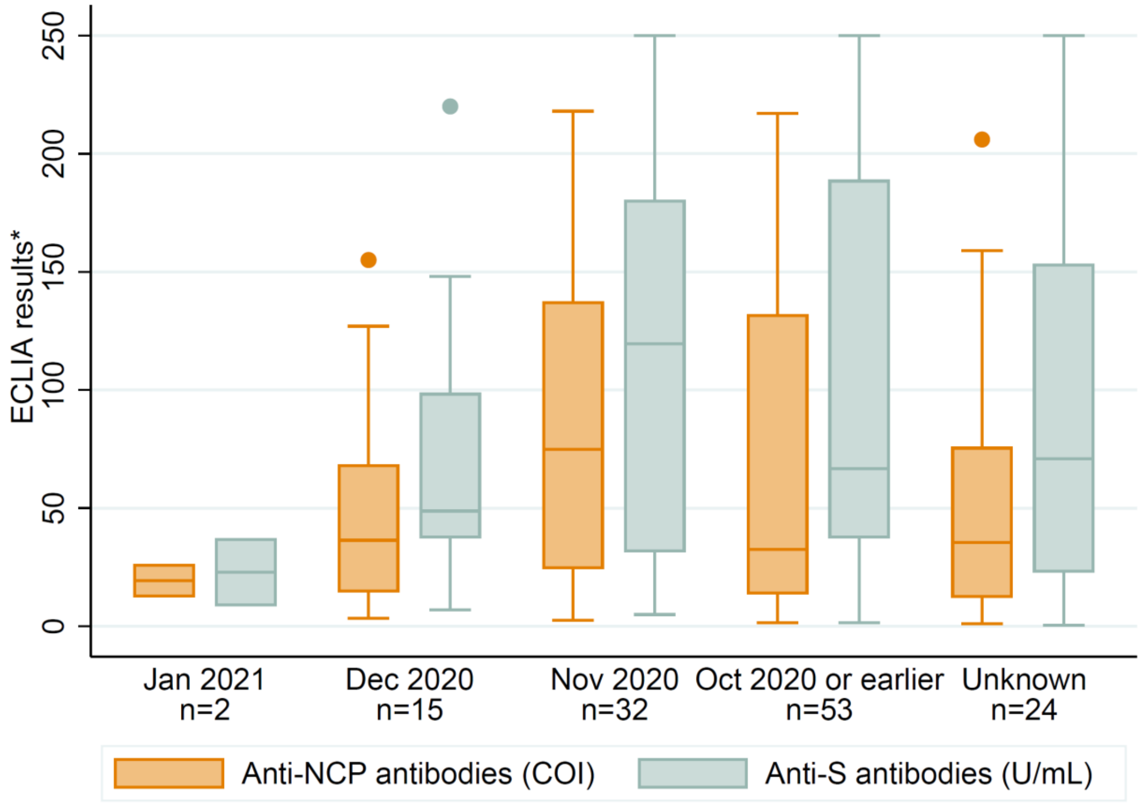


COI = cut-off index. Units are presented according to the manufacturer's recommendations.
99 study participants reported the time point of a positive nasopharyngeal swab test, 3 individuals reported the time point of COVID-19 (without a test but several symptoms consistent with the diseases), and 24 study participants did not know a time point ("unknown").

**Supplemental figure C:** Random forest algorithm to identify specific symptoms that separated seropositive and negative participants.

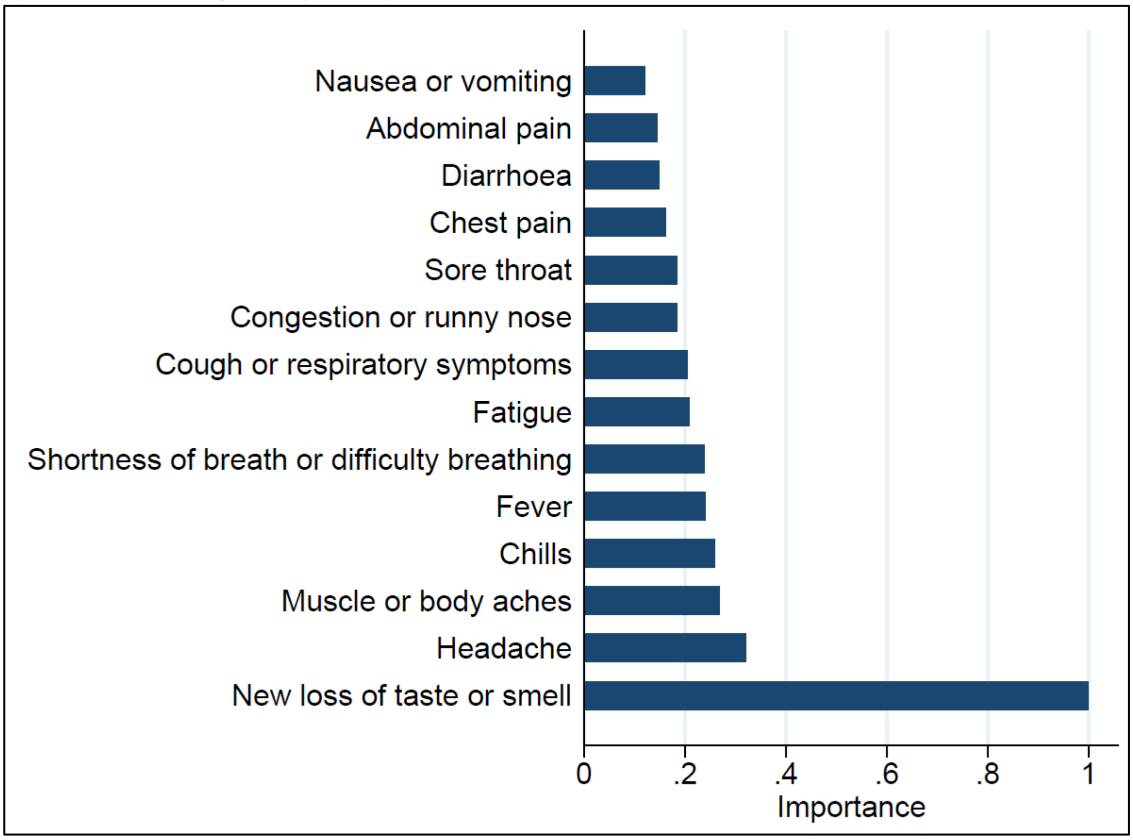


The symptom that separated seropositive and negative participants best was "new loss of taste or smell."

**Supplemental table D:** Comparison between seropositive and seronegative individuals categorized according to the use of personal protective equipment (PPE) and hygiene precautions.

| PPE and hygiene precautions | Responses (N=972) | Study participants (N=978) | Seropositive (N=126) | Seronegative (N=852) | P |
|---|---|---|---|---|---|
| Masks | 978 | 975 (100%) | 126 (100%) | 849 (100%) | 1.00 |
| Gloves | 838 | 315 (32%) | 45 (36%) | 270 (32%) | 0.36 |
| Goggles or safety glasses | 773 | 68 (7.0%) | 5 (4.0%) | 63 (7.4%) | 0.19 |
| Face shields | 759 | 22 (2.2%) | 4 (3.2%) | 18 (2.1%) | 0.51 |
| Disinfectants | 972 | 956 (98%) | 126 (100%) | 830 (97%) | 0.10 |
| Others | 630 | 41 (4.2%) | 1 (0.79%) | 40 (4.7%) | 0.05 |
| ≥3 of the above | 978 | 361 (37%) | 49 (39%) | 312 (37%) | 0.62 |

**Supplemental table E:** Subjective assessment (questionnaire) of the source of transmission in seropositive study participants (N=126).

| Questions and answers on the source of transmission of SARS-CoV-2 | N Responses | Seropositive (N=126) |
|---|---|---|
| **Do you know the source of transmission?** | 125 | |
| No | | 23 (18%) |
| Yes, I am certain. | | 65 (52%) |
| Probably (likely yes) | | 10 (7.9%) |
| Maybe, possibly | | 11 (8.7%) |
| Uncertain (likely no) | | 16 (13%) |
| **Time point of transmission?** | 102 | |
| Last month (January 2021) | | 2 (1.6%) |
| Two months ago (December 2020) | | 15 (12%) |
| Three months ago (November 2020) | | 32 (25%) |
| Four months or more ago | | 53 (42%) |
| **Place/Location of transmission?** | 102 | |
| During working hours: in the office/at the police station | | 15 (12%) |
| During working hours: fieldwork outside the police station | | 18 (14%) |
| Outside working hours: contact within the family/in the household | | 48 (38%) |
| Outside working hours: contact outside the family/household contacts | | 16 (13%) |
| Other | | 5 (4.0%) |
| **If you selected "during working hours," please indicate the (presumed) contact situation.** | 33 | |
| Next to a colleague who was not wearing a mask* | | 5 (4.0%) |
| Desk sharing | | 1 (0.79%) |
| Lunch break/eating break | | 1 (0.79%) |
| In the police car | | 6 (4.8%) |
| Arrest/detention of a suspect | | 4 (3.2%) |
| Police operation during demonstration, contact with a crowd | | 2 (1.6%) |
| Other | | 14 (11%) |
| **If you selected "outside working hours," please indicate the (presumed) contact situation.** | 64 | |
| At home/within the same household | | 38 (30%) |
| Outside home: family reunion | | 10 (7.9%) |
| Meeting friends, going out, sports activity | | 9 (7.1%) |
| Public transport, shopping | | 2 (1.6%) |
| Outside Switzerland | | 1 (0.79%) |
| Other | | 4 (3.2%) |

*Wearing face masks for employees of the Bern Cantonal Police was recommended on August 28, 2020, and made mandatory on October 13, 2020. The questionnaire addressed situations since the onset of the pandemic in Switzerland (February 25, 2020).

**Supplemental table F:** Spearman's rank correlation coefficients between serum antibody titres of anti-NCP (1) and anti-S antibodies (2) and serum neutralization titres towards SARS-CoV-2 (3, 4, 5).

| Variables | (1) | (2) | (3) | (4) | (5) |
|---|---|---|---|---|---|
| (1) Anti-NCP antibodies (COI) | 1.000 | | | | |
| (2) Anti-S1 antibodies (U/mL) | 0.550 | 1.000 | | | |
| (3) D614G | 0.527 | 0.675 | 1.000 | | |
| (4) B.1.1.7 (alpha) | 0.532 | 0.687 | 0.861 | 1.000 | |
| (5) B.1.351 (beta) | 0.363 | 0.567 | 0.644 | 0.647 | 1.000 |

All corresponding P values are <0.001.

**Supplemental figure D:** Antibody titres (y-axis) of seropositive study participants in association with dilution titres (x-axis) in neutralization assays against isogenic SARS-CoV-2 viruses harbouring the D614G spike (N=126).

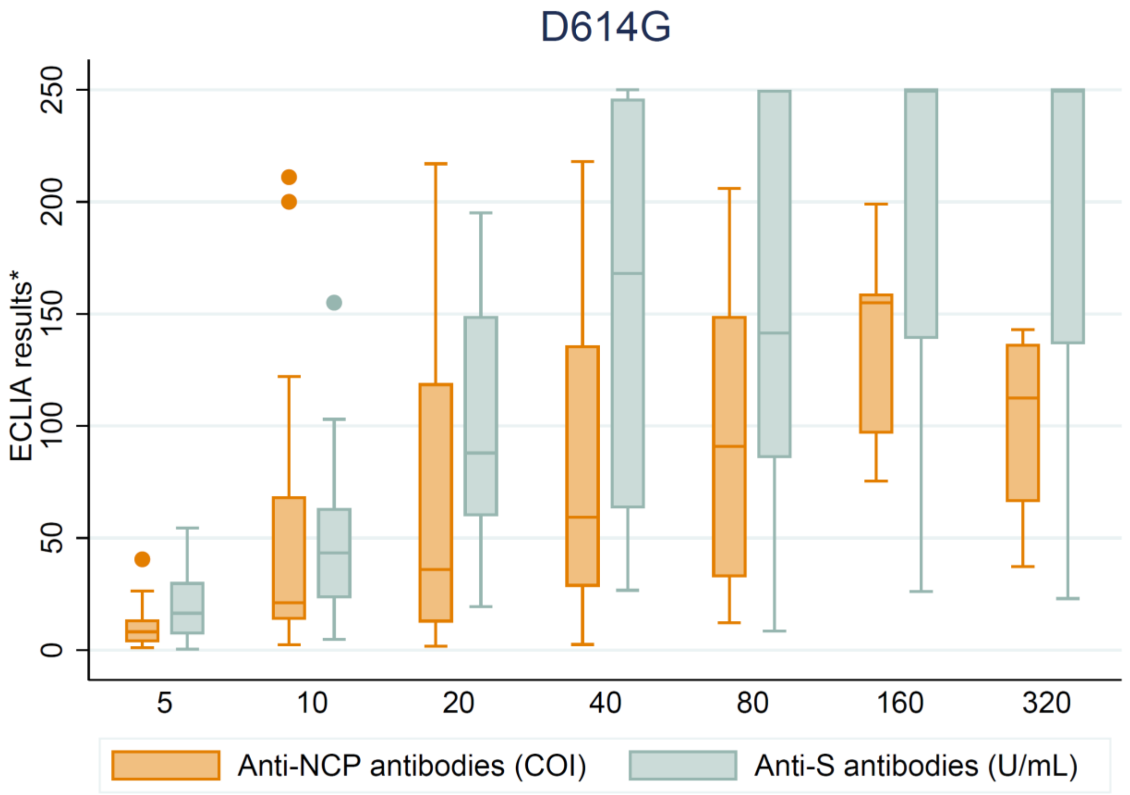


*The units are presented according to the manufacturer's recommendations. COI = cut-off index.

**Supplemental figure E:** Antibody titres (y-axis) of seropositive study participants in association with dilution titres (x-axis) in neutralization assays against isogenic SARS-CoV-2 harbouring the full-length B.1.1.7 spike (N=126).

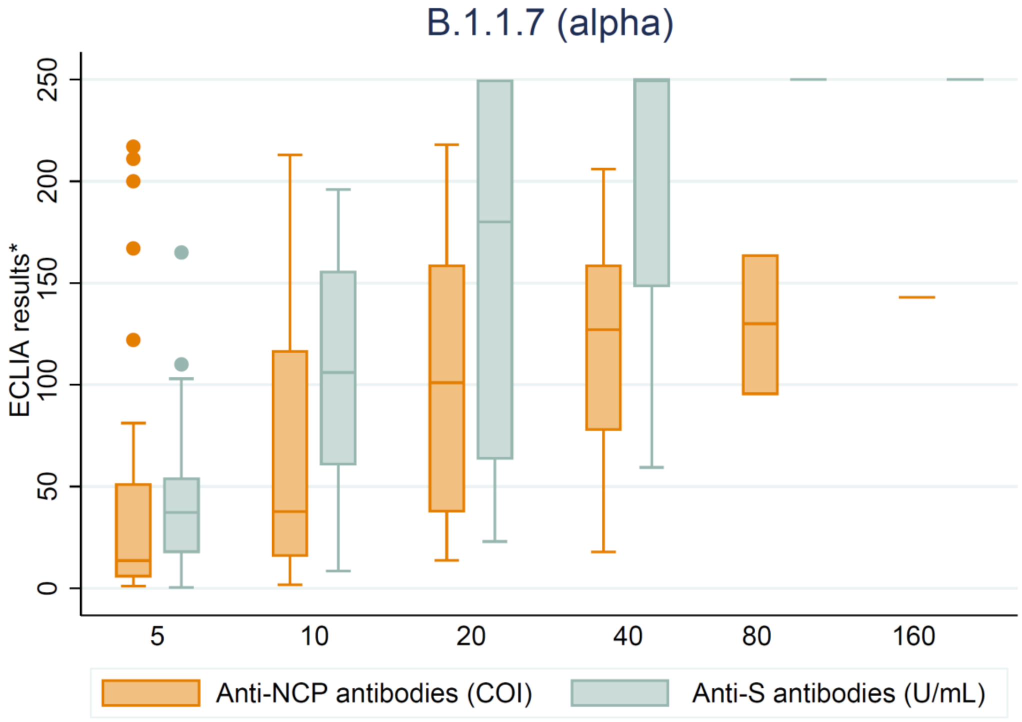


*The units are presented according to the manufacturer's recommendations; (N=126). COI = cut-off index.

**Supplemental figure F:** Antibody titres (y-axis) of seropositive study participants in association with dilution titres (x-axis) in neutralization assays against isogenic SARS-CoV-2 viruses harbouring the full-length B.1.351 spike (N=126).

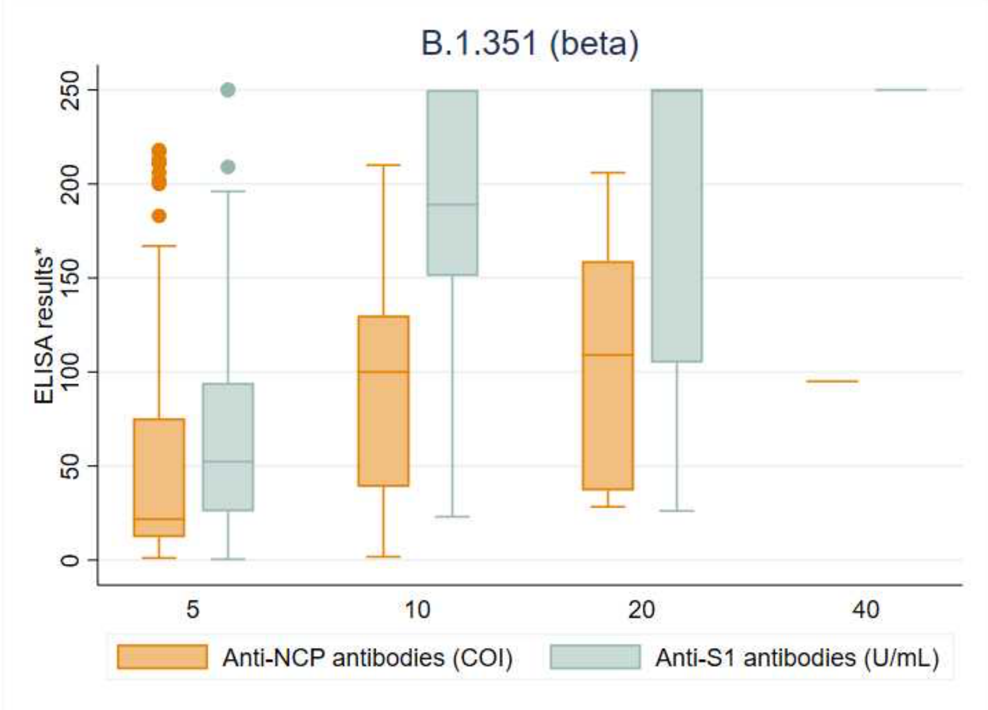


*The units are presented according to the manufacturer's recommendations. COI = cut-off index.

**Supplemental table G:** Association of antibody titres and serum neutralization titres.

| Serum neutralization titres | Covariate antibody titres | OR (95% CI) | P |
|---|---|---|---|
| Assays with D614G | anti-NCP titres (continuous) | 1.008 (1.003 to 1.014) | 0.003 |
| | anti-NCP>37.5 | 3.1 (1.5 to 6.1) | 0.001 |
| | anti-S1 titres (continuous) | 1.019 (1.013 to 1.024) | <0.001 |
| | anti-S>65 | 6.7 (3.2 to 14.3) | <0.001 |
| **Serum neutralization titres** | **Covariate antibody titres** | **OR (95% CI)** | **P** |
| Assays with spike from B.1.1.7 (alpha) | anti-NCP titres (continuous) | 1.008 (1.002 to 1.014) | 0.007 |
| | anti-NCP>37.5 | 2.9 (1.4 to 6.2) | 0.005 |
| | anti-S1 titres (continuous) | 1.021 (1.015 to 1.027) | <0.001 |
| | anti-S>65 | 6.7 (3.1 to 14.7) | <0.001 |
| **Serum neutralization titres** | **Covariate antibody titres** | **OR (95% CI)** | **P** |
| Assays with spike from B.1.351 (beta) | anti-NCP titres (continuous) | 1.002 (0.995 to 1.010) | 0.559 |
| | anti-NCP>37.5 | 3.0 (1.1 to 8.1) | 0.030 |
| | anti-S1 titres (continuous) | 1.021 (1.014 to 1.028) | <0.001 |
| | anti-S>65 | 6.1 (2.1 to 17.9) | 0.001 |

Note that level of serum neutralization titres is modelled as ordered categories because a normal distribution of error terms cannot be assumed; hence, linear regression is not suitable and median regression does not seem adequate, given that dependent variables are discrete values on the log scale.

Anti-NCP antibody titres above the median (>37.5 COI) were associated with about a three-fold increased level of neutralization, whereas anti-S1 antibody titres above the median (>65 U/mL) were associated with about a six-fold increase. The association of antibody titres as a continuous variable was also more pronounced for anti-S1 antibody titres than for anti-NCP antibody titres with odds ratios per unit close to 1.
